# Extraction of clinical information from faxed medical records using a small local large language model pipeline on consumer hardware

**DOI:** 10.64898/2026.10.03.26364667

**Authors:** Zhouwen Tang

## Abstract

**Objective:** Specialty referral packets often arrive by fax and clinicians must manually read and organize a large amount of disparate information to affect continuity of care. Commercial large language models (LLMs) can organize packet information, but protected health information (PHI) must be secured. We evaluated whether an open-weight pipeline running entirely on one consumer graphics card can accurately extract clinical information from faxed referral packets.

**Materials and Methods:** We generated 200 synthetic colorectal cancer referral packets from simulated patient records, rendered them as faxes without a text layer. A local pipeline transcribed each packet with an optical character recognition model (OvisOCR2) and extracted structured data with an open-weight LLM (Qwen3.8-27B). Twenty outcomes spanning patient identifiers, laboratory results, dates, diagnoses, procedures, medications, imaging, and event chronology were scored against ground truth on 140 validation packets.

**Results:** Laboratory values were extracted exactly in 98.9% of 7879 instances, dates in 99.7%, diagnoses in 100%, and procedures in 92.9%. Timeline recall was 96.7% and precision 99.4%. Weighted accuracy was 0.990 (95% CI 0.987–0.992). None of 11,993 extracted facts were fabricated. Median processing time was 90 seconds per packet.

**Conclusion:** An open-weight pipeline on consumer hardware accurately extracted multi-domain clinical information from synthetic faxed referrals.

## BACKGROUND AND SIGNIFICANCE

Despite the pervasiveness of electronic health records, health care communications between practices and systems often relies on telefacsimile (“fax”) due to interoperability concerns between different systems, low cost of implementation, reliability of transmission, and familiarity with established methods ^1^. One example of such communications is the ambulatory patient handoff in the form of specialty referrals. Such transitions in care are challenging because the receiving provider must synthesize complex patient information domains such as diagnoses, labs, imaging, medications, and procedures to build a temporal narrative of the patient’s previous care. Adding to the challenge, faxes, even when inserted into the electronic health record (EHR), are flat digital media devoid of text layers that can be easily extracted to interact with the EHR’s native information search and retrieval capabilities. The receiving healthcare provider must then comb through pages of unfamiliar clinical information presented in an unfamiliar and disjointed format for relevant clinical information.

Large language models (LLM) excel at reading a bounded document and extracting information from its contents ^2,3^ and have been studied as aids for clinical documentation ^4,5^. Indeed, general purpose proprietary or “frontier” models often have superior performance for medical knowledge and clinical queries when compared to healthcare specific models ^6^. As a result, commercial AI platforms have sprung up to provide services for both consumer and enterprise healthcare customers ^7^. However, privacy legislation such as the General Data Protection Regulation (GDPR) in Europe and Health Insurance Portability and Accountability Act in the United States limit sharing protected health information (PHI) with outside entities and a business associate agreement, institutional review, and an ongoing trust relationship remain significant barriers to many practices.

Small open-weight LLMs democratize access to frontier capabilities at a fraction of the cost by allowing local customization and deployment ^8^. Local deployment also sequesters PHI within the HIPAA covered entity, avoiding business associate agreements and avoiding uploading PHI to the broader internet or outside servers.

We built a medical record ingestion pipeline to summarize incoming new patient referral information using open weight LLMs that is deployed locally on inexpensive consumer grade hardware. We then evaluated the accuracy of this pipeline with synthetic patient data.

## OBJECTIVE

To determine whether an entirely local, open-weight optical character recognition and language-model pipeline can extract clinically relevant information (laboratory values, procedures, diagnoses, medications, and event chronology) from faxed gastroenterology referral packets with accuracy sufficient for triage; and to construct a shareable benchmark, with clinically audited ground truth, for evaluating such pipelines.

## MATERIALS AND METHODS

### Validation and test set construction

A set of synthetic patient referral packets were fabricated using data generated with Synthea, an open-source patient simulator ^9^. Packets were scanned for usable colorectal cancer referral arcs. Colorectal cancer arcs were selected because this disease state is associated with a plausible set of labs, imaging, and procedures leading to referral for specialty care.

Each patient extracted was assigned to a bounded window ending at the referral date, producing a structured record consisting of clinical encounters, diagnoses, procedures, imaging, laboratory results with units and dates, and medications. This structured record served as the ground truth for comparison. Clinical prose in the form of referral letter, procedure reports, imaging report, and laboratory result reports were generated using a proprietary LLM (Claude Sonnet 5; Anthropic, PBC; San Francisco, CA, USA) under a verification gate that confirmed every ground-truth value appeared verbatim in the rendered text and that no additional clinically scoreable value had been introduced.

Documents were then rendered to fax format with 1728×2155 pixels per page, 1-bit depth, and Consultative Committee for International Telegraphy and Telephony (CCITT) Group 4 compression. Packets did not contain a text layer. Visual presentation was further randomized across fonts styles and sizes, letterhead layouts, table styles, and document order. Each packet also carried one or two distractor passages drawn from the patient’s own unrelated comorbidities (Supplemental Figure 1). The packets were then split into 60 packet development set and a 140 packet validation set.

### Medical record ingestion pipeline construction

The pipeline was built using OvisOCR2 ^10^ served through vLLM (Inferact; San Francisco, CA, USA) for optical character recognition (OCR) and extraction was built with Qwen3.8-27B ^11^ quantized to Nvidia Floating-Point 4-bit (NVFP4) and served through NInfer inference engine ^12^. A 120,000 token context was specified so that the entire packet may be read at once rather than retrieved in fragments. Both models fit concurrently on a single NVIDIA RTX 5090 graphical processing unit (Figure 1).

**Figure 1.**
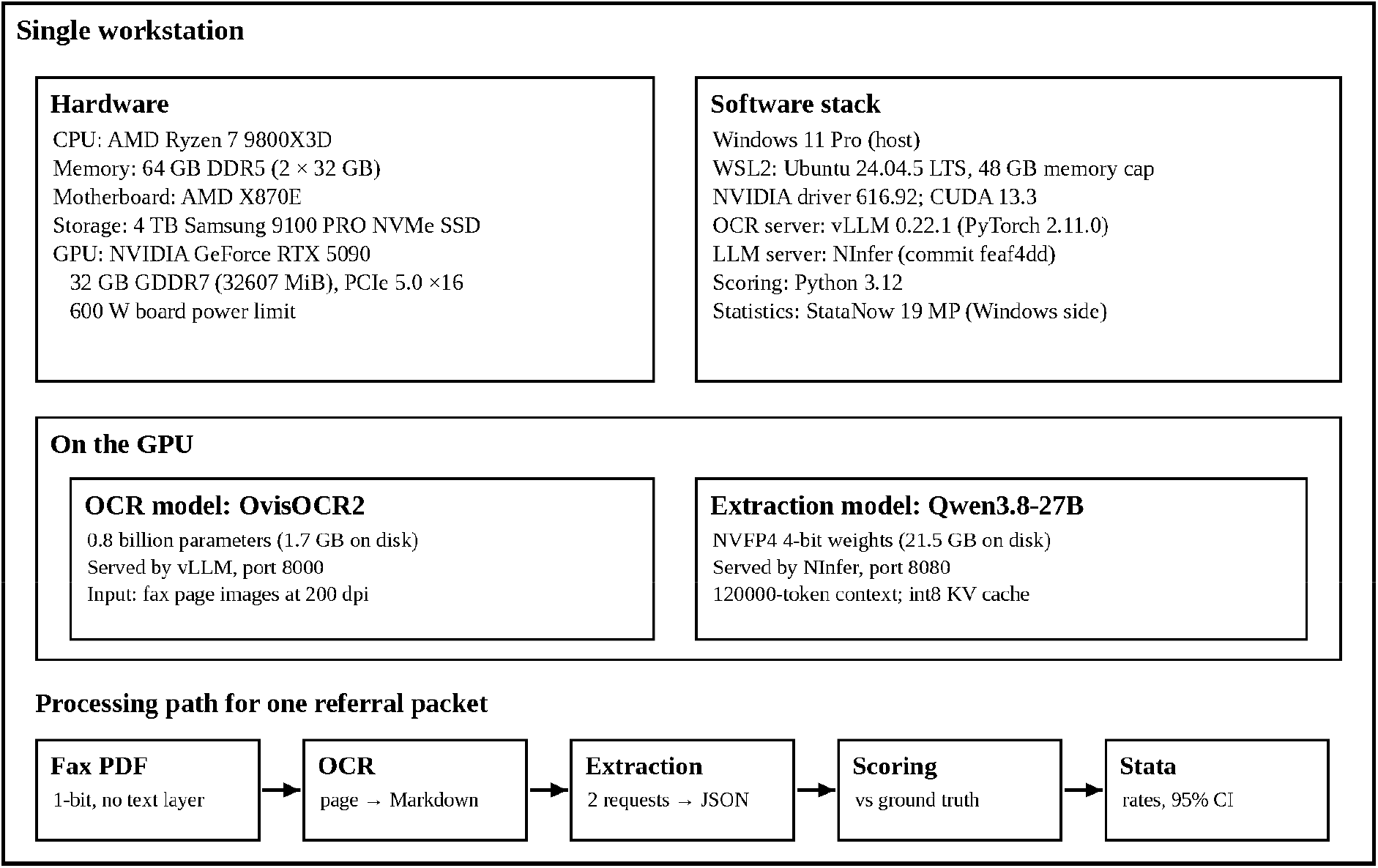
Pipeline hardware and software specifications.

Extraction was performed with two requests per packet, each given the complete transcribed packet. The first request returned patient identifiers, diagnoses, procedures, medications, imaging, and the event chronology, with the model’s reasoning mode enabled with reasoning level set to medium. The second returned laboratory results and the most-recent-value list, with reasoning disabled. The two outputs were merged before scoring.

The extraction prompt (Supplemental Appendix A) specified the model to answer only from the packet, distinguish not documented from documented as negative, cite the source page of every fact, and transcribe values without rounding or conversion. Extraction was output to structured JavaScript Object Notation (JSON).

### Outcomes

Twenty pre-specified data field outcomes were scored with a binary judgement when compared to the ground truth.

1. **Patient identifier outcomes** of matching on name, date of birth, and medical record number.
2. **Laboratory outcomes** of captured boolean, and value exact and unit correct.
3. **Dates outcomes** of both exact day, month, and year as well as only month and year, pooled across every dated fact.
4. **Diagnosis and procedure outcomes** of captured boolean.
5. **Medication outcomes** of captured boolean and dose correct.
6. **Imaging outcomes** of captured boolean and exact date match.
7. **Chronology outcomes** consisting of a comparison with the reference ground truth timeline which included every dated diagnosis, procedure, imaging study, and one laboratory collection per collection date. Medications were excluded from events for the chronology outcome. For chronology outcomes, recall is the proportion of reference events matched by an extracted timeline entry. Precision is the proportion of extracted entries that matched a reference event in the timeline. Timeline ordering was summarized by Kendall’s tau-b between reference and extracted sequence positions of one-to-one matched events.
8. **Recency outcomes** consisted of each outcome measured more than once in a packet and whether the model returned the most recent value, the correct date, and both jointly.

Other outcomes included hallucination, contradiction, and documented negatives parameters. A hallucination is defined as an extracted item matching no ground-truth, distractor, or authored content. A contradiction is a documented diagnosis recorded as negative or not done. Finally, a single weighted aggregate summarizes every outcome. This score was computed per packet as the weighted mean of that packet’s field rates under fixed clinical-consequence weights, then averaged across packets. (Supplemental Table S1).

## Statistical analysis

Each packet’s extraction was compared with its ground truth by a scoring program (Python 3.12, Python Software Foundation, Wilmington, DE, USA). Extracted and reference items were first paired one-to-one. Each paired item was then judged on each applicable outcome. Values were compared exactly, units were compared after normalizing equivalent notations, and dates after conversion to calendar dates so that date format differences were not penalized. Chronology entries were paired with reference events by the same name rule and by date to the month, under the rules described above.

Scored outcomes were exported to Stata (StataNow 19 MP; StataCorp, College Station, TX) as two datasets: a long file with one record per field instance, carrying the packet identifier, the outcome, and the binary result; and a packet-level file with one record per packet carrying that packet’s weighted aggregate, chronology F1, and tau-b. A packet’s laboratory and recency outcomes were included only if its laboratory request parsed, and its remaining outcomes only if its clinical request parsed, a rule specified before the validation run. All 140 clinical requests and 139 of 140 laboratory requests parsed, so 140 packets contribute to every outcome except the seven laboratory and recency outcomes, which rest on 139.

Each outcome was estimated as the pooled proportion of correct instances over ground-truth instances across packets. Wilson score intervals were computed for reference. Because instances within a packet share a single generated document set, page layout, and transcription, they are not independent. Primary 95% confidence intervals were therefore obtained by nonparametric cluster bootstrap with the packet as the resampling unit. Packets were drawn with replacement. The weighted aggregate, equal-weight aggregate, and mean chronology F1 were bootstrapped over the 140 packet-level values by the same procedure.

## RESULTS

Weighted accuracy across the 20 outcomes was 0.990 (95% CI 0.987–0.992). Equal weight across all outcomes was 0.990 (0.988–0.992).

### Synthetic referral packet construction

Of 39,927 simulated patients, 285 (0.71%) carried the trigger diagnosis of colorectal cancer and 263 (92.3% of trigger-positive) produced a usable arc. The remainder of arcs were rejected for having no colonoscopy (n=13) or fewer than eight laboratory results (n=9). The first 200 usable arcs in order were built into packets (Table 1).

**Table 1.** Referral packet characteristics.

| Characteristic | Value |
| --- | --- |
| Simulated patients scanned | 39,927 |
| Trigger-positive (colorectal cancer) | 285 (0.71%) |
| Usable referral arcs | 263 (92.3% of trigger-positive) |
| Packets generated | 200 (60 development / 140 validation) |
| Pages per packet, median (range) | 9 (6–19) |
| Ground-truth facts per packet, median (range) | 65 (20–262) |
| Documents per packet, median (range) | 7 (5–7) |
| Fax format | 1728×2155 px, 1-bit, CCITT G4, no text layer |
| Visual variants | 6 typefaces, 5 sizes, 7 letterheads, 4 table styles, 189 document orders |
| Packets with distractor passages | 166 (83%) |

### OCR transcription

One thousand nine hundred and thirty-five pages were generated across 200 packets. Numeric-token and date-token recall were 1.000 in every packet, and word recall was below 1.000 in only two packets by one token each. In one packet, an uncommon given name was normalized to its common spelling (Elizbeth read as Elizabeth). In the other packet, the salt of doxycycline hyclate was corrupted to “doxycycline hycline.” Transcription took a median of 11.0 seconds per packet with a median of 1.24 seconds per page.

### Completeness of extraction

All 280 extraction requests (two per packet) were completed with a normal stop. Every clinical request was completed. However, a single request out of 140 laboratory requests produced an unparseable string. Under the rule specified before the run, that packet contributes to every outcome except the laboratory and recency outcomes.

### Field extraction accuracy

Laboratory values were transcribed exactly in 98.9% of 7879 instances, with units correct in 99.2% and 99.6% of analytes captured. Dates were correct in 99.7% of 9931 instances. Of the 25 date misses, 15 were laboratory dates and 10 procedure dates. Diagnoses were captured 100% of 1,363 instances and procedures in 92.9% of 742 instances. Of the 53 missed procedures, 50 were colonoscopies and polypectomies and 3 were colon biopsies; 44 of the 53 are mentioned in the history line of a later endoscopy report (“prior colonoscopy on 04/08/2006”). In those cases the model recorded the colonoscopy that report described, but not the earlier one it cites. Patient name, date of birth, and medical record number were correct in every packet. Table 2 gives the full results for the 140 validation packets.

**Table 2.** Extraction accuracy outcomes.

| Field | k/n | Rate | Wilson 95% CI | Clustered 95% CI | Mean number per packet |
| --- | --- | --- | --- | --- | --- |
| Patient name | 140/140 | 1.000 | 0.973–1.000 | 1.000–1.000 | 1.0 |
| Patient date of birth | 140/140 | 1.000 | 0.973–1.000 | 1.000–1.000 | 1.0 |
| Patient record number | 140/140 | 1.000 | 0.973–1.000 | 1.000–1.000 | 1.0 |
| Laboratory captured | 7879/7913 | 0.996 | 0.994–0.997 | 0.994–0.997 | 56.9 |
| Laboratory value, exact | 7793/7879 | 0.989 | 0.987–0.991 | 0.984–0.994 | 56.7 |
| Laboratory unit correct | 7818/7879 | 0.992 | 0.990–0.994 | 0.987–0.997 | 56.7 |
| Date exact | 9906/9931 | 0.997 | 0.996–0.998 | 0.996–0.999 | 70.9 |
| Date, month and year | 9907/9931 | 0.998 | 0.996–0.998 | 0.996–0.999 | 70.9 |
| Diagnosis captured | 1363/1363 | 1.000 | 0.997–1.000 | 1.000–1.000 | 9.7 |
| Procedure captured | 689/742 | 0.929 | 0.908–0.945 | 0.909–0.947 | 5.3 |
| Medication captured * | 735/735 | 1.000 | 0.995–1.000 | 1.000–1.000 | 5.7 |
| Medication dose correct * | 735/735 | 1.000 | 0.995–1.000 | 1.000–1.000 | 5.7 |
| Imaging captured * | 140/140 | 1.000 | 0.973–1.000 | 1.000–1.000 | 1.0 |
| Imaging date exact * | 140/140 | 1.000 | 0.973–1.000 | 1.000–1.000 | 1.0 |
| Chronology recall | 2957/3058 | 0.967 | 0.960–0.973 | 0.954–0.977 | 21.8 |
| diagnoses | 1337/1363 | 0.981 | 0.972–0.987 | 0.964–0.993 | 9.7 |
| procedures | 697/742 | 0.939 | 0.920–0.954 | 0.918–0.958 | 5.3 |
| imaging | 292/298 | 0.980 | 0.957–0.991 | 0.943–1.000 | 2.1 |
| laboratory collections | 631/655 | 0.963 | 0.946–0.975 | 0.947–0.978 | 4.7 |
| Chronology precision | 2782/2799 | 0.994 | 0.990–0.996 | 0.991–0.997 | 20.0 |
| Chronology, event dating | 2954/2957 | 0.999 | 0.997–1.000 | 0.998–1.000 | 21.1 |
| Chronology, ordering (tau-b) | — | 0.956 | — | — | — |
| Most-recent value | 2018/2128 | 0.948 | 0.938–0.957 | 0.913–0.975 | 16.1 |
| Most-recent date | 2125/2128 | 0.999 | 0.996–1.000 | 0.997–1.000 | 16.1 |
| Most-recent value + date | 2018/2128 | 0.948 | 0.938–0.957 | 0.913–0.975 | 16.1 |
| Weighted aggregate | — | 0.990 | — | 0.987–0.992 | — |

### Timeline extraction accuracy

Three thousand and fifty-eight events were eligible for timeline construction. Of these, chronology recall was 96.7%. Events were correctly dated in 99.9%, and ordering was closely correlated with the ground truth (Kendall tau-b 0.956). Recall by domain was 98.1% for diagnoses, 93.9% for procedures, 98% for imaging, and 96.3% for laboratory collections.

Of the 101 events not recalled, 3.3% of the eligible events, 45 were missing procedures. These included 23 procedures referenced only in the history line of a later endoscopy report and 20 procedures on dates the timeline covered with other entries (for example, the polyp diagnosis or stool test) but without a separate entry for the procedure itself; two fit neither pattern. Most of the remainder were diagnoses, labs, and imaging listed on a single line when the matching algorithm could only count one event per line. For example, a single entry reading “respiratory distress, hypoxemia, pneumonia, COVID-19…” counted as one match for nine reference diagnoses, even though the extraction’s diagnosis list contained all nine. Three laboratory collections were recorded under wording the scorer did not recognize (for example, “prostate specific Ag collected”).

The model wrote a total of 3716 timeline events. Of these, 75 were additional entries for a laboratory-draw day already counted, 676 were events excluded from the ground truth reference (eye exams, therapy, workflow events), and 166 were extractions of distractor events inserted into the packets. This yielded 2799 events scored for precision of which 2782 matched a ground truth event yielding a precision of 99.4%. Seventeen extracted events were not judged to have a ground truth match, but all were extractions of information elsewhere in the packet such as referrals, medication stops, and wellness visits. These extractions reflected real information in packets but their ground truth references were not scored. None were hallucinations.

### Fabrication

Of 11,993 extracted facts, the scorer flagged three as not matching any ground truth. On examination of these possible fabrications, all three were present in the packet: two were diagnoses (type 2 diabetes mellitus and essential hypertension) that originated in the LLM generating the packet and not in Synthea and were thus not in the ground truth, and one was “hot snare polypectomy” correctly extracted from the endoscopy reports but reported as a procedure distinct from the ground truth polypectomy. Therefore, there were no fabrications stemming from the pipeline.

In addition, no contradictions and no decimal-shift or digit-transposition errors were found in the 7879 laboratory values.

Distractor content was pulled into structured output at 8.6% (125 of 1,446 opportunities): 34 of 60 distractor diagnoses, 30 of 245 immunizations, 58 of 651 procedures, and 3 of 490 unrelated vital signs.

### Performance and cost

Median end-to-end extraction was 80.3 seconds per packet, 52.3 seconds for the clinical request and 27 seconds for the laboratory request, at an effective 221 output tokens per second and transcription added 11 seconds per packet. The validation run consumed 2.03 million input and 2.91 million output tokens (Table 3).

**Table 3.** Pipeline technical performance.

| Measure | Median | Max |
| --- | --- | --- |
| Extraction, s/packet (both requests) | 80.3 | 208.3 |
| clinical request, s | 52.3 | 101.4 |
| laboratory request, s | 27.0 | 121.0 |
| Transcription, s/packet | 11.0 | 28.6 |
| Transcription, s/page | 1.24 | 1.91 |
| Output tokens/packet (both requests) | 17,651 | 47,098 |
| clinical request | 11,197 | 22,086 |
| laboratory request | 6,198 | 27,846 |
| Input tokens per request | 6,700 | 14,460 |
| Effective decode rate, output tokens/s | 221 | — |
| Token volume, validation run | 2.03 M input / 2.91 M output | — |

## DISCUSSION

We created a medical record ingestion pipeline with an OCR model linked with a general small local LLM. The pipeline runs entirely on local consumer hardware. We then created synthetic patient specialty referral packets in fax format and demonstrated via a validation set that pipeline extraction of diagnosis, procedure, imaging, medication, and chronology domains from these packets were highly accurate. By using a highly tuned model specific inference engine (NInfer) for our extraction model, we were able to maximize general speed given the limitations of our hardware.

Though the function and accuracy of this pipeline is likely easily matched by large frontier models, the advantage of this pipeline is rooted in its local deployment. By not depending on externally hosted tools or application programming interface (API) calls, PHI can reliably remain confidential while avoiding potentially complex legal and administrative burdens of business associate agreements for HIPAA compliance.

There are several important limitations to this study. Primarily, synthetic packets are imperfect mimics of real-world referral packets. In our study, packets only cover one clinical archetype. While colorectal cancer referrals tend to naturally involve communications of a wide range of clinical information domains, the performance of the pipeline on this single archetype should not be generalized to broader clinical situations that may differ in the content and makeup of referral packets. More broadly, while care was taken to vary the format and page order of referral packets, real word packets are expected to be more heterogeneous and the essentially perfect OCR performance in this study may degrade.

Though our model did not exhibit hallucinatory output on our validation set, this outcome cannot be generalized to real world use. Indeed, the model prompt was specifically written to limit model extraction of extraneous information. The intent of this pipeline is simply to organize disparate referral packet information. Clinical judgement must lie with the physician user. The model must be specifically prompted to not insert clinical judgement in production.

Additionally, this study only tests one concurrent user. Production deployments may expect several concurrent users and deployment on current hardware may lead to performance degradation or further limitations in context.

Further work will be needed in generalizing the performance of this pipeline on a wide range of real-world referral packets compared with human extracted ground truth variables, including with multiple concurrent users and on more capable hardware.

## Data Availability

All data produced in the present study are available upon reasonable request to the authors.

