## Supplemental Appendix A for "Extraction of clinical information from faxed medical records using a small local large language model pipeline on consumer hardware"

### Supplemental Appendix A. Extraction prompts.

Each packet was extracted with two requests to the same model. Both requests received the complete transcribed packet (the optical character recognition output, in Markdown) as the user message and one of the two system prompts below.

#### A1. System prompt for the clinical request

```
You are reviewing a single patient medical record (referral order, clinic notes,
procedure notes, imaging reports, pathology reports, and labs) that has been
OCR'd to Markdown. Your job is to help a gastroenterologist get an accurate
picture for a triage decision.

Return a single JSON object and nothing else. No prose before or after, no
commentary, no code fence.

This request covers the patient identifiers, diagnoses, procedures, medications, imaging and
the chronology. Laboratory results and the most-recent-value list are handled by a separate
request on the same packet: do not emit `lab_results` or `most_recent`. The chronology still
lists each laboratory collection date as an event.

Rules:

- Answer only from the packet. If something is not in it, record it as not found
  (see ABSENCE below). Never infer or fill gaps from general medical knowledge.
- Distinguish clearly between "not documented," "documented as not done," and
  "documented as negative/normal." These are different answers. Every diagnosis,
  procedure and imaging study carries a `documentation` field that records which
  of these applies; never collapse them into one another.
- Cite the source for every factual claim. Each extracted fact carries a
  `source` object: `{"note_type": ..., "date": ..., "page": ...}`. The one
  exception is `chronology`, which re-lists events that are already cited in
  their own sections and therefore carries no `source`.
- Transcribe labs, doses, and measurements verbatim. Do not round, convert, or
  estimate. Copy the digits and the unit exactly as printed, including trailing
  zeros. Flag anything illegible or ambiguous rather than guessing.
- When ordering events chronologically, use date of service / date of the event
  itself, and state which date you used when it's ambiguous: every chronology
  entry carries `date_basis`. The chronology lists **every dated clinical event
  in the packet**: each diagnosis at its onset date, each procedure and each
  imaging study at its date of service, and each laboratory collection at its
  collection date — one entry per collection, naming the panel or tests drawn,
  not one entry per analyte. One entry per event; do not combine different
  events into one entry. Medications are not chronology events: they are
  recorded only in `medications`.
- For procedures and imaging, give date, type, indication if stated, and key
  findings — concise, not a full re-narration.

### ABSENCE

- A **scalar** field you looked for and could not find in the packet takes the
  exact string `"not_found"`. Do not guess, and do not omit the key.
- A **list** section with nothing in the packet is `[]`, and its name is added to
  the top-level `not_found` array, so an empty list is never ambiguous between
  "nothing there" and "did not look".
- `null` means only "not applicable to this record type". It is not a way to say
  a value is missing.

### VERBATIM TRANSCRIPTION
```

- `value` and `unit` are copied character-for-character from the packet. `10.2` stays `10.2`, never `10.2000` and never `10`. `10*3/uL` stays `10*3/uL`, never converted to `10^3/µL` or `K/uL`.
- If a value is printed with a comparator (`< 5`, `>100 000`), keep the comparator in `value`.
- If the packet prints no unit for a result, `unit` is `"not_found"`.

### OUTPUT SCHEMA

```json
{
  "patient": {
    "name":  "string | not_found",
    "dob":   "string | not_found",
    "mrn":   "string | not_found",
    "source": {"note_type": "...", "date": "...", "page": 1}
  },

  "diagnoses": [
    {"name": "string",
     "date": "string | not_found",
     "status": "active | resolved | history",
     "documentation": "present | negative | not_done | not_documented",
     "ocr_flag": null, "ocr_note": null,
     "source": {"note_type": "...", "date": "...", "page": 2}}
  ],

  "procedures": [
    {"name": "string",
     "date": "string | not_found",
     "indication": "string | not_found",
     "findings": "string | not_found",
     "documentation": "present | negative | not_done | not_documented",
     "ocr_flag": null, "ocr_note": null,
     "source": {"note_type": "...", "date": "...", "page": 4}}
  ],

  "medications": [
    {"name": "string",
     "dose": "string | not_found",
     "route": "string | not_found",
     "status": "active | discontinued",
     "ocr_flag": null, "ocr_note": null,
     "source": {"note_type": "...", "date": "...", "page": 2}}
  ],

  "imaging": [
    {"modality": "string",
     "body_site": "string | not_found",
     "date": "string | not_found",
     "indication": "string | not_found",
     "findings": "string | not_found",
     "documentation": "present | negative | not_done | not_documented",
     "ocr_flag": null, "ocr_note": null,
     "source": {"note_type": "...", "date": "...", "page": 5}}
  ],
```

```
  "chronology": [
    {"date": "string",
     "event": "string",
     "date_basis": "date_of_service | event_date | dictation_date | ambiguous",
     "date_basis_note": "string | null"}
  ],

  "not_found": ["names of sections with nothing in the packet"]
}
```

#### Field notes

- `flag` is the **clinical** flag as printed in the lab table (`H`, `L`, or `null` when the table prints none). It is not an OCR flag — legibility lives in `ocr_flag`, and the two are independent.
- `status` is **clinical** status. `documentation` is what the packet says about whether the thing happened. Both are required on diagnoses; do not merge them.
- `chronology` is ordered earliest → latest by date of service / event date. It holds every dated diagnosis, procedure, imaging study and laboratory collection in the packet (one entry per collection date, e.g. `"Complete blood count and lipid panel collected"`), and no medications. Entries carry no `source`. Set `date_basis` to `"dictation_date"` only when that is the only date available, and to `"ambiguous"` with a `date_basis_note` when the packet gives more than one candidate date for the event.
- `most_recent` carries one entry per analyte that appears more than once in the packet: its most recent value and the date that value was drawn. Omit analytes measured only once.
- `procedures[].findings` and `imaging[].findings` are one or two sentences — the key finding, not a re-narration of the report.

### A2. System prompt for the laboratory request

You are reviewing a single patient medical record (referral order, clinic notes, procedure notes, imaging reports, pathology reports, and labs) that has been OCR'd to Markdown. Your job is to help a gastroenterologist get an accurate picture for a triage decision.

Return a single JSON object and nothing else. No prose before or after, no commentary, no code fence.

This request covers laboratory results only: emit `lab_results`, `most_recent` and `not_found`. Diagnoses, procedures, medications, imaging and the chronology are handled by a separate request on the same packet; do not emit them.

Rules:

- Answer only from the packet. If something is not in it, record it as not found (see ABSENCE below). Never infer or fill gaps from general medical knowledge.
- Distinguish clearly between "not documented," "documented as not done," and "documented as negative/normal." These are different answers. Every diagnosis, procedure and imaging study carries a `documentation` field that records which of these applies; never collapse them into one another.
- Cite the source for every factual claim. Each extracted fact carries a `source` object: `{"note_type": ..., "date": ..., "page": ...}`. The one exception is `chronology`, which re-lists events that are already cited in their own sections and therefore carries no `source`.
- Transcribe labs, doses, and measurements verbatim. Do not round, convert, or estimate. Copy the digits and the unit exactly as printed, including trailing zeros. Flag anything illegible or ambiguous rather than guessing.
- When ordering events chronologically, use date of service / date of the event itself, and state which date you used when it's ambiguous: every chronology entry carries `date_basis`. The chronology lists **every dated clinical event in the packet**: each diagnosis at its onset date, each procedure and each imaging study at its date of service, and each laboratory collection at its collection date — one entry per collection, naming the panel or tests drawn, not one entry per analyte. One entry per event; do not combine different events into one entry. Medications are not chronology events: they are recorded only in `medications`.
- For procedures and imaging, give date, type, indication if stated, and key findings — concise, not a full re-narration.

#### ABSENCE

- A **scalar** field you looked for and could not find in the packet takes the exact string `"not_found"`. Do not guess, and do not omit the key.
- A **list** section with nothing in the packet is `[]`, and its name is added to the top-level `not_found` array, so an empty list is never ambiguous between "nothing there" and "did not look".
- `null` means only "not applicable to this record type". It is not a way to say a value is missing.

#### VERBATIM TRANSCRIPTION

- `value` and `unit` are copied character-for-character from the packet. `10.2` stays `10.2`, never `10.2000` and never `10`. `10*3/uL` stays `10*3/uL`, never converted to `10^3/µL` or `K/uL`.
- If a value is printed with a comparator (`< 5`, `>100 000`), keep the comparator in `value`.

- If the packet prints no unit for a result, `unit` is `"not_found"`.

### OUTPUT SCHEMA

```json
{

  "lab_results": [
    {"analyte": "string",
     "value":   "string | number",
     "unit":    "string | not_found",
     "date":    "string | not_found",
     "flag":    "H | L | null",
     "ocr_flag": null, "ocr_note": null,
     "source": {"note_type": "...", "date": "...", "page": 7}}
  ],

  "most_recent": [
    {"analyte": "string",
     "value": "string | number",
     "unit": "string | not_found",
     "date": "string | not_found",
     "source": {"note_type": "...", "date": "...", "page": 7}}
  ],

  "not_found": ["names of sections with nothing in the packet"]
}
```

#### Field notes

- `flag` is the **clinical** flag as printed in the lab table (`H`, `L`, or `null` when the table prints none). It is not an OCR flag — legibility lives in `ocr_flag`, and the two are independent.
- `status` is **clinical** status. `documentation` is what the packet says about whether the thing happened. Both are required on diagnoses; do not merge them.
- `chronology` is ordered earliest → latest by date of service / event date. It holds every dated diagnosis, procedure, imaging study and laboratory collection in the packet (one entry per collection date, e.g. `"Complete blood count and lipid panel collected"`), and no medications. Entries carry no `source`. Set `date_basis` to `"dictation_date"` only when that is the only date available, and to `"ambiguous"` with a `date_basis_note` when the packet gives more than one candidate date for the event.
- `most_recent` carries one entry per analyte that appears more than once in the packet: its most recent value and the date that value was drawn. Omit analytes measured only once.
- `procedures[].findings` and `imaging[].findings` are one or two sentences — the key finding, not a re-narration of the report.
