## Supplemental Figure 1 for "Extraction of clinical information from faxed medical records using a small local large language model pipeline on consumer hardware"

**Supplemental Figure 1. Sample synthetic referral packet as received by the pipeline.**

Nine pages of a sample synthetic colorectal cancer referral packet as generated from Synthea patient arc. Pages appear in randomized document order. The ground truth for this packet holds 21 laboratory results, 7 diagnoses, 12 procedure rows, 4 medications, and 15 distractor facts.

#### North Shore Physicians Group — Primary Care

104 Endicott Street, Danvers, MA 01923

#### FACSIMILE TRANSMITTAL

**TO:** New Patient Coordinator, Gastroenterology — Section of Gastroenterology & Hepatology

**FROM:** Peter K. Lindqvist, DO, Internal Medicine
**DATE:** 31-Dec-2017
**PAGES (including this cover):** 9

**RE:** Gillian Hettinger · DOB 30-Sep-1966 · MRN 8352971

[ X ] For review and scheduling   [  ] Urgent   [  ] Please call to confirm receipt

**Message:**

Please find enclosed a referral for Gillian Hettinger for evaluation of a newly diagnosed colonic malignancy. Enclosed documents: referral letter, two colonoscopy / pathology reports, and a laboratory report. Kindly contact our office at 978-555-0188 to arrange an appointment.

*CONFIDENTIALITY NOTICE: This facsimile is intended only for the use of the individual or entity to which it is addressed and may contain information that is privileged and confidential. If you are not the intended recipient, any disclosure, copying, or distribution is prohibited. If you have received this facsimile in error, please notify us by telephone and return the original to us by mail.*

### Letter of Referral

**Patient:** Gillian Hettinger **DOB:** 30-Sep-1966 (Age 51, F) **MRN:** 8352971 **Date of Referral:** 31-Dec-2017

#### REASON FOR REFERRAL

Referring patient with newly diagnosed Adenocarcinoma of the rectosigmoid colon (onset 31-Dec-2017) for specialist evaluation and management.

#### ALLERGIES

No known drug allergies.

#### HISTORY OF PRESENT ILLNESS

Ms. Hettinger is a 51-year-old woman presenting with iron deficiency and fatigue, with no overt rectal bleeding. She was found to have Adenocarcinoma of the rectosigmoid colon, onset 31-Dec-2017. Her history is notable for recurrent rectosigmoid polyp and colonic polyp (rectosigmoid), as detailed below.

#### MEDICATIONS

Recently discontinued:

- { (Dienogest Estradiol Valerate ) (Dienogest Estradiol Valerate ) (Estradiol Valerate ) (Estradiol Valerate ) (Inert Ingredients ) } Pack 2 mg PO (discontinued 28-Jun-2012)
- Etonogestrel Drug Implant 68 mg (discontinued 23-Jun-2013)
- { (Ethinyl Estradiol Norgestimate ) (Ethinyl Estradiol Norgestimate ) (Ethinyl Estradiol Norgestimate ) (Inert Ingredients ) } Pack 0.035 mg PO (discontinued 13-Jun-2015)
- { (Ethinyl Estradiol Levonorgestrel ) (Inert Ingredients ) } Pack 0.03 mg PO (discontinued 02-Jun-2017)

#### PAST MEDICAL HISTORY

**Problem List:**

- Adenocarcinoma of the rectosigmoid colon, onset 31-Dec-2017 — newly diagnosed
- Recurrent rectosigmoid polyp, onset 24-Dec-2017
- Colonic polyp (rectosigmoid), onset 07-Oct-2016
- History of miscarriage, onset 08-May-2006
- Chronic pain, onset 02-Feb-2014
- Chronic low back pain, onset 02-Feb-2014
- Fibromyalgia, onset 02-Feb-2014

**Relevant Procedures:**

- Implantation of subcutaneous contraceptive, 28-Jun-2012
- Movement therapy, 27-Feb-2014
- Movement therapy, 02-Mar-2014
- Movement therapy, 05-Mar-2014
- Implantation of subcutaneous contraceptive, 08-Jul-2016
- Removal of subcutaneous contraceptive, 08-Jul-2016
- Removal of subcutaneous contraceptive, 16-Jun-2017

#### ASSESSMENT AND PLAN

Newly diagnosed Adenocarcinoma of the rectosigmoid colon in the setting of iron deficiency and fatigue, without overt rectal bleeding, and a recent history of recurrent rectosigmoid polyp and colonic polyp (rectosigmoid). Referral placed for specialist assessment and coordination of oncologic management. Please evaluate and advise on further workup and treatment planning.

### North Shore Physicians Group — Primary Care — Clinical Laboratory

104 Endicott Street, Danvers, MA 01923

#### LABORATORY REPORT

| | |
|---|---|
| Patient | Gillian Hettinger |
| DOB | 30-Sep-1966 |
| MRN | 8352971 |

##### Complete Blood Count

Collected: 19-Oct-2012 Reported: 19-Oct-2012

| Test | Result | Units |
|---|---|---|
| WBC | 8 | 10*3/uL |
| RBC | 5.4 | 10*6/uL |
| Hemoglobin | 15.5 | g/dL |
| Hematocrit | 49.7 | % |
| MCV | 94.3 | fL |
| MCH | 27.3 | pg |
| MCHC | 34.5 | g/dL |
| RDW-SD | 45.1 | fL |
| Platelet Count | 172.5 | 10*3/uL |
| MPV | 10.3 | fL |
| PDW | 14.6 | fL |
| Cholesterol, Total | 206.3 | mg/dL |
| LDL Cholesterol | 125 | mg/dL |
| HDL Cholesterol | 53.5 | mg/dL |
| Triglycerides | 138.5 | mg/dL |

##### Laboratory Studies

Collected: 02-Feb-2014 Reported: 02-Feb-2014

| Test | Result | Units |
|---|---|---|
| Drugs of abuse 5 panel - Urine by Screen method | Negative (qualifier value) | n/a |

##### Lipid Panel

Collected: 30-Sep-2016 Reported: 30-Sep-2016

| Test | Result | Units |
|---|---|---|
| Cholesterol, Total | 159.5 | mg/dL |
| LDL Cholesterol | 87.9 | mg/dL |

| Test | Result | Units |
|---|---|---|
| HDL Cholesterol | 49.4 | mg/dL |
| Triglycerides | 111.2 | mg/dL |

Reference ranges are laboratory-specific and provided for interpretation only. L = below reference range, H = above.

### North Shore Physicians Group — Primary Care

104 Endicott Street, Danvers, MA 01923

#### INTERVAL PRIMARY CARE NOTE

*Enclosed for completeness. Not related to the reason for referral.*

##### Vital Signs — Wellness Visit 06-Oct-2017

- Height: 165.3 cm
- Weight: 76.7 kg
- Body mass index: 28.1 kg/m2
- Systolic blood pressure: 113 mmHg
- Diastolic blood pressure: 78 mmHg
- Heart rate: 91 /min
- Respiratory rate: 14 /min

Vitals recorded by nursing staff at the routine annual visit; reviewed and unchanged from the prior year.

##### Health Maintenance

Immunizations on file:

- Influenza split virus trivalent PF, administered 30-Sep-2016
- zoster vaccine live, administered 06-Oct-2017
- Influenza split virus trivalent PF, administered 06-Oct-2017
- Td (adult) 5 Lf tetanus toxoid preservative free adsorbed, administered 06-Oct-2017

Preventive screening completed 06-Oct-2017: Assessment of health and social care needs; Assessment of anxiety; Assessment of substance use; Screening for drug abuse.

All screening instruments were negative and no further action was indicated at that visit.

### Riverbend Medical Imaging

Department of Radiology · Diagnostic Imaging Report

**Patient:** Gillian Hettinger
**DOB:** 30-Sep-1966 **MRN:** 8352971
**Date of Service:** 27-Dec-2017
**Ordering Provider:** Peter K. Lindqvist, DO, North Shore Physicians Group — Primary Care
**Accession:** CT165378

#### EXAMINATION

CT of the chest, abdomen and pelvis with intravenous and oral contrast.

#### INDICATION

Biopsy-proven adenocarcinoma of the rectosigmoid colon. Staging prior to surgical referral.

#### TECHNIQUE

Helical acquisition from the lung apices through the ischial tuberosities following administration of intravenous iodinated contrast. Oral contrast was given. Multiplanar reformations were reviewed.

#### COMPARISON

None available.

#### FINDINGS

**Chest:** No pulmonary nodule or mass. No pleural effusion. No mediastinal or hilar lymphadenopathy. Heart size normal.

**Abdomen:** Liver is normal in size and contour with no focal hepatic lesion. Gallbladder, pancreas, spleen, adrenal glands and kidneys are unremarkable. No ascites. No retroperitoneal lymphadenopathy.

**Pelvis:** Circumferential wall thickening of the rectosigmoid colon over a segment of approximately 4.2 cm, with mild stranding of the adjacent pericolonic fat. Two adjacent mesorectal lymph nodes measure up to 9 mm in short axis. No pelvic free fluid. Bladder is unremarkable.

**Bones:** No aggressive osseous lesion.

#### IMPRESSION

1. Circumferential rectosigmoid wall thickening with mild pericolonic stranding, consistent with the known biopsy-proven primary. Radiologic stage at least T3.
2. Two borderline mesorectal nodes up to 9 mm; indeterminate, cannot exclude nodal involvement.
3. No evidence of hepatic or pulmonary metastatic disease.

Electronically signed by the interpreting radiologist.

### Colonoscopy & Polypectomy Report

**Date of Procedure:** 07-Oct-2016

#### INDICATION

Screening for occult blood in feces, reason: Polyp of colon. Fecal immunoassay (Hemoglobin.gastrointestinal.lower [Presence] in Stool by Immunoassay --1st specimen) obtained 07-Oct-2016: 12.7 ng/mL.

#### PREPARATION

The preparation was adequate (Boston Bowel Preparation Scale 8/9).

#### PROCEDURE

Colonoscopy performed 07-Oct-2016. The colonoscope was advanced to the cecum; the terminal ileum was not intubated. Cecal landmarks (appendiceal orifice, ileocecal valve) were identified.

#### FINDINGS

A single 8 mm sessile polyp was identified in the rectosigmoid colon at approximately 18 cm. The remainder of the examined colon was normal. No mass, no active bleeding, no diverticulosis.

#### INTERVENTION

Rectal polypectomy performed, reason: Polyp of colon. The polyp was removed in a single piece by hot snare polypectomy. Estimated blood loss was minimal. There were no immediate complications.

#### PATHOLOGY

Pathology addendum: rectosigmoid polyp — tubular adenoma with low-grade dysplasia; margin free of adenoma.

#### DIAGNOSIS

Colonic polyp (rectosigmoid), onset 07-Oct-2016, active.

#### IMPRESSION

Rectosigmoid tubular adenoma with low-grade dysplasia, status post complete snare polypectomy on 07-Oct-2016.

### Colonoscopy & Polypectomy Report

**Date of Procedure:** 24-Dec-2017

**Indication/Diagnosis:** Recurrent rectosigmoid polyp (onset 24-Dec-2017, active).

**Procedures Performed:** Colonoscopy; Rectal polypectomy (reason: Recurrent rectal polyp).

#### PREPARATION

The preparation was adequate (Boston Bowel Preparation Scale 7/9).

#### EXTENT OF EXAMINATION

The colonoscope was advanced to the cecum. Cecal landmarks were identified and photodocumented.

#### FINDINGS

At the prior rectosigmoid polypectomy site (approximately 18 cm) there was a recurrent 12 mm sessile polyp with a firm, partially fixed base. Additional biopsies were taken from the polyp base and the adjacent mucosa.

#### TECHNIQUE

The recurrent polyp was removed piecemeal by snare. Hemostasis was achieved with two endoscopic clips. Estimated blood loss was minimal.

#### PATHOLOGY ADDENDUM

Pathology addendum: rectosigmoid polyp and base biopsies — invasive adenocarcinoma arising in a tubulovillous adenoma; adenocarcinoma extends to the cauterized deep margin.

#### ASSESSMENT

Recurrent rectosigmoid polyp, status post polypectomy, with pathology as above. Active diagnosis; referral for oncologic/surgical evaluation indicated.
