## Supplemental Table S1 for "Extraction of clinical information from faxed medical records using a small local large language model pipeline on consumer hardware"

### Supplemental Table S1. Outcome weights

| Domain | Outcome | Weight |
|---|---|---|
| Identifiers | Patient name | 2 |
| Identifiers | Patient date of birth | 2 |
| Identifiers | Patient record number | 3 |
| Laboratory | Laboratory captured | 3 |
| Laboratory | Laboratory value, exact | 3 |
| Laboratory | Laboratory unit correct | 3 |
| Dates | Date exact | 2 |
| Dates | Date, month and year | 1 |
| Diagnosis | Diagnosis captured | 3 |
| Procedure | Procedure captured | 2 |
| Medication | Medication captured | 2 |
| Medication | Medication dose correct | 3 |
| Imaging | Imaging captured | 1 |
| Imaging | Imaging date exact | 1 |
| Chronology | Chronology recall | 1 |
| Chronology | Chronology precision | 1 |
| Chronology | Chronology, event dating | 1 |
| Recency | Most-recent value | 2 |
| Recency | Most-recent date | 2 |
| Recency | Most-recent value + date | 3 |

Each of the 20 outcomes was assigned an weight of 1, 2, or 3 according to the clinical consequence of an error for referral triage. A weighted mean of each packet's outcome rates were calculated. Overall weighted accuracy is the mean of the packet scores. The 95% confidence interval is the 2.5th to 97.5th percentile of 5000 bootstrap resamples of packets.
